# Individuals with recent prior SARS-CoV-2 infection are at reduced risk of Omicron infection and associated hospitalization

**DOI:** 10.1101/2022.08.10.22278641

**Authors:** Mihika Nadig, Michiel JM Niesen, Patrick Lenehan, Vineet Agarwal, Jason Ross, Sankar Ardhanari, AJ Venkatakrishnan, Venky Soundararajan

## Abstract

Omicron sub-lineages such as BA2.12.1 and BA5 have breached prior infection-induced immunity and vaccine-induced immunity. This capacity of Omicron to reinfect patients calls for a characterization of vaccination, infection, and reinfection patterns. We analyzed de-identified longitudinal electronic health records for 389,746 individuals (88,679 fully-vaccinated, 184,205 boosted, 73,184 with prior infection) across multiple US states. Compared to individuals with only full vaccination, the rates of SARS-CoV-2 infections in the Omicron era were reduced for individuals with additional prior infection (1.4 to 1.8-fold reduced, depending on vaccine status) or booster vaccination (1.3 to 2.0-fold reduced). Although prior infection was associated with lower incidence of SARS-CoV-2 infection, we found that the relative risk (RR) of infections for individuals with prior infection has increased during Omicron. During October, 2021, RR was 0.11 [0.10-0.13, 95% CI] while during May, 2022, it increased to 0.57 [0.46-0.68, 95% CI], suggesting an increase in reinfections with Omicron. Furthermore, we found that time since prior infection is associated with risk of reinfection, providing evidence of waning immunity. Prior infections before June, 2021, were associated with marginal reduction in risk of infection (eg., RR = 0.80 [0.68-0.90] for prior infection during January, 2021), while recent prior infections were associated with significant reduction in risk (eg., RR = 0.24 [0.20-0.29, 95% CI] for prior infection during November, 2021). Despite an observed increase in reinfections and vaccine breakthrough infections, our findings emphasize the protective effect of natural and vaccine immunity, with prior infection providing ∼6 months of protection from reinfection.

## Introduction

Nearly 550 million SARS-CoV-2 cases have been recorded globally since the beginning of the COVID-19 pandemic (as of July 1, 2022)^1^. At a population level there has been continual altering of the host’s immunological constitution triggered by SARS-CoV-2 infections and reciprocally there has been continual evolution of the virus’s antigenic constitution under the host’s immune selection pressure.^2–4^ As a result, new immuno-evasive variants such as the Delta variant and the Omicron variant have emerged and driven surges in new infections.^5,6^ On the other side, hosts have acquired infection-induced immunity, vaccine-induced immunity or both — “hybrid immunity” (**Figure 1a**). Over 11.7 billion vaccine doses have been administered globally^7^. The Omicron variant has broken through immunity induced by prior infections, vaccines (including boosters), and both in several individuals. This may in part be aided by the waning effectiveness of natural immunity and the vaccines.^8–11^ The ongoing surge of infections caused by Omicron, and its sub-lineages such as BA.5, have thus highlighted the gaps in our understanding of the risks of breakthrough infections and their severity.

**Figure 1:**
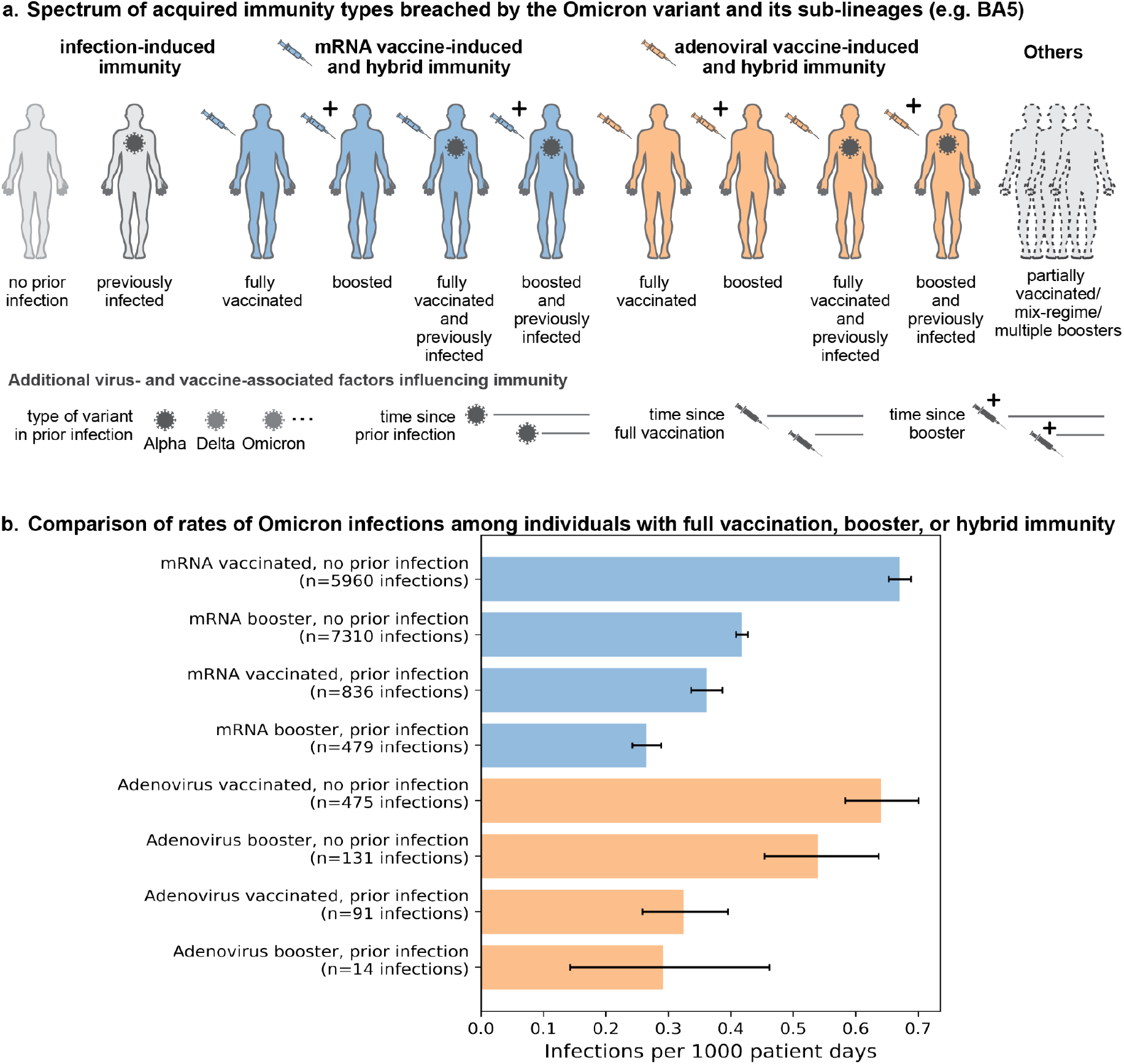
Infections during Omicron (December 2021 - June 2022), stratified by patient vaccine and prior infection status. **(a)** Schematic depiction of the types of acquired immunity included in this study. **(b)** Rates of infection, normalized by eligible patient days, during a time-period of high Omicron prevalence (December 1st, 2021 - May 29, 2022) for individuals with specific types of previously acquired immunity against SARS-CoV-2. Error bars indicate 95% confidence intervals.

Understanding infection risk and disease severity requires analysis of longitudinal clinical data from the real-world setting. To this end, we have analyzed de-identified electronic health record data from over 380,000 individuals from health systems spread across the US, through the nference Analytics Platform.^12^ Here, we characterize patient cohorts based on infection-induced and vaccine-induced immunity and compare the rates of breakthrough infections and re-infections during the Omicron era. We also compare the severity of the infections through the analyses of hospitalization events in these cohorts. Finally, we highlight the need for such a conceptual framework in order to help guide health policy decisions on future rollouts of vaccines.

## Results

In order to characterize the incidence and severity of Omicron infections and identify the risks of reinfection with Omicron, we retrospectively analyzed reinfections in US health systems, spread across multiple states, based on analysis of fully de-identified and HIPAA compliant electronic health records.

### Rate of Omicron infections in patients with prior infection and vaccination is lower than in patients with vaccination only

We first evaluated the risk of SARS-CoV-2 infection during a time of high Omicron prevalence (December 1, 2021 -May 29, 2022). Risk of infection is quantified as the number of infections per 1000 eligible patient days, for cohorts of patients (**Figure 1a, Table 1-2**) stratified by their known COVID-19 vaccine status and their prior SARS-CoV-2 infection status at the study start date (December 1, 2021). We found that individuals with prior infection and booster vaccination had reduced incidence of SARS-CoV-2 infection during Omicron, compared with individuals that were fully vaccinated without booster or prior infection (**Figure 1b**). Patients with a previous SARS-CoV-2 infection had lower incidence rates of infection (i.e. infections per 1000 patient days) during the Omicron era than patients who were not previously infected for each assessed vaccination status (mRNA vaccinated: 0.36 [0.34-0.39, 95% CI] versus 0.67 [0.65-0.69, 95% CI]; mRNA boosted: 0.26 [0.24-0.29, 95% CI] versus 0.42 [0.41-0.43, 95% CI]; adenovirus vaccinated: 0.32 [0.26-0.40, 95% CI] versus 0.64 [0.58-0.70, 95% CI]; and adenovirus boosted: 0.29 [0.14-0.46, 95% CI] versus 0.54 [0.45-0.64, 95% CI]). A similar reduction in risk of infection in individuals with prior exposure to SARS-CoV-2 was observed for individuals with unknown vaccine status (**Figure S1**).

**Table 1:**
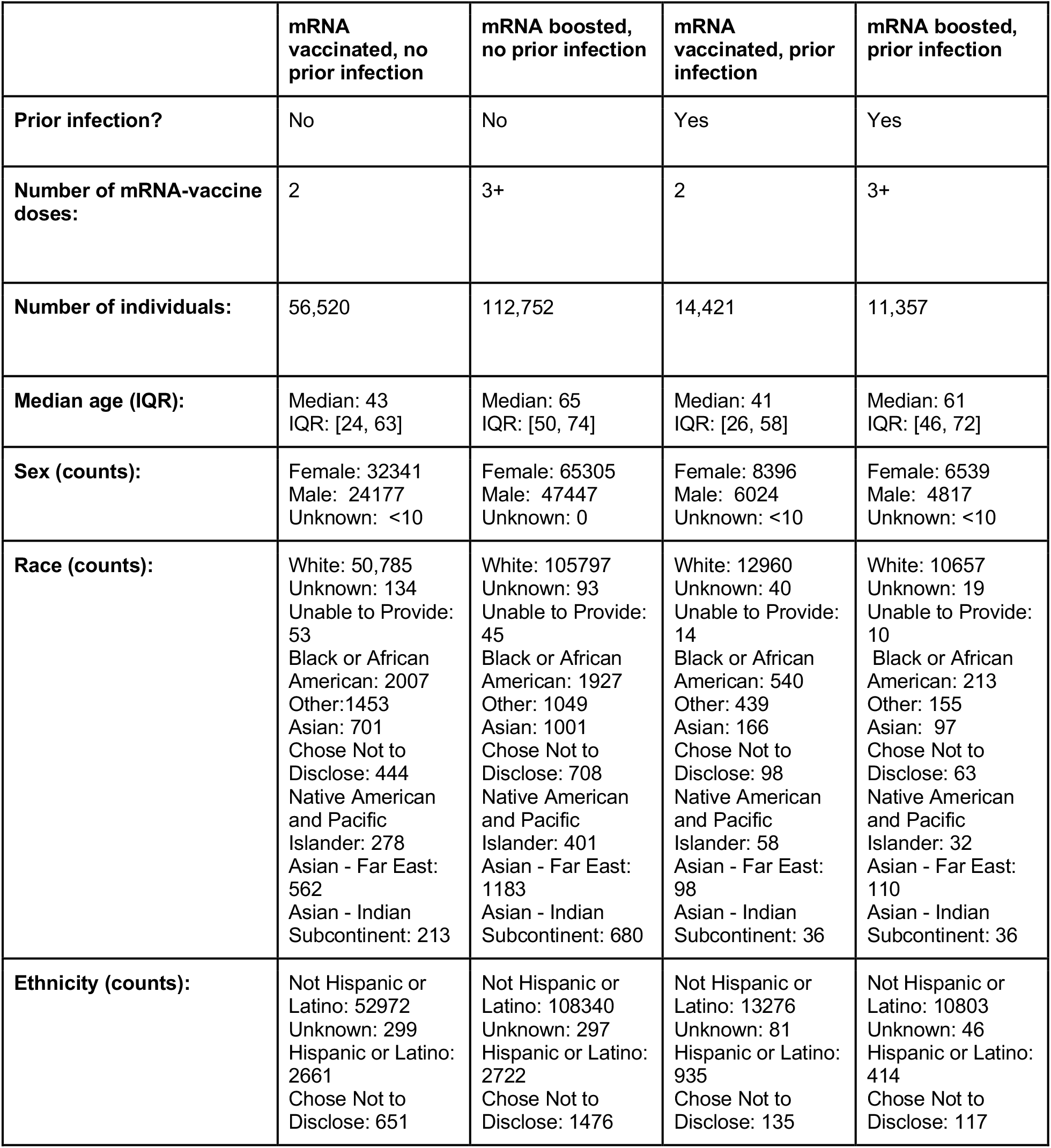
Cohort characteristics for mRNA-vaccinated individuals.

|  | mRNA vaccinated, no prior infection | mRNA boosted, no prior infection | mRNA vaccinated, prior infection | mRNA boosted, prior infection |
| --- | --- | --- | --- | --- |
| **Prior infection?** | No | No | Yes | Yes |
| **Number of mRNA-vaccine doses:** | 2 | 3+ | 2 | 3+ |
| **Number of individuals:** | 56,520 | 112,752 | 14,421 | 11,357 |
| **Median age (IQR):** | Median: 43<br>IQR: [24, 63] | Median: 65<br>IQR: [50, 74] | Median: 41<br>IQR: [26, 58] | Median: 61<br>IQR: [46, 72] |
| **Sex (counts):** | Female: 32341<br>Male: 24177<br>Unknown: <10 | Female: 65305<br>Male: 47447<br>Unknown: 0 | Female: 8396<br>Male: 6024<br>Unknown: <10 | Female: 6539<br>Male: 4817<br>Unknown: <10 |
| **Race (counts):** | White: 50,785<br>Unknown: 134<br>Unable to Provide: 53<br>Black or African American: 2007<br>Other: 1453<br>Asian: 701<br>Chose Not to Disclose: 444<br>Native American and Pacific Islander: 278<br>Asian - Far East: 562<br>Asian - Indian Subcontinent: 213 | White: 105797<br>Unknown: 93<br>Unable to Provide: 45<br>Black or African American: 1927<br>Other: 1049<br>Asian: 1001<br>Chose Not to Disclose: 708<br>Native American and Pacific Islander: 401<br>Asian - Far East: 1183<br>Asian - Indian Subcontinent: 680 | White: 12960<br>Unknown: 40<br>Unable to Provide: 14<br>Black or African American: 540<br>Other: 439<br>Asian: 166<br>Chose Not to Disclose: 98<br>Native American and Pacific Islander: 58<br>Asian - Far East: 98<br>Asian - Indian Subcontinent: 36 | White: 10657<br>Unknown: 19<br>Unable to Provide: 10<br>Black or African American: 213<br>Other: 155<br>Asian: 97<br>Chose Not to Disclose: 63<br>Native American and Pacific Islander: 32<br>Asian - Far East: 110<br>Asian - Indian Subcontinent: 36 |
| **Ethnicity (counts):** | Not Hispanic or Latino: 52972<br>Unknown: 299<br>Hispanic or Latino: 2661<br>Chose Not to Disclose: 651 | Not Hispanic or Latino: 108340<br>Unknown: 297<br>Hispanic or Latino: 2722<br>Chose Not to Disclose: 1476 | Not Hispanic or Latino: 13276<br>Unknown: 81<br>Hispanic or Latino: 935<br>Chose Not to Disclose: 135 | Not Hispanic or Latino: 10803<br>Unknown: 46<br>Hispanic or Latino: 414<br>Chose Not to Disclose: 117 |

**Table 2:** Cohort characteristics for adenovirus-vaccinated individuals.

|  | **Adenovirus vaccinated, no prior infection** | **Adenovirus boosted, no prior infection** | **Adenovirus vaccinated, prior infection** | **Adenovirus boosted, prior infection** |
| --- | --- | --- | --- | --- |
| **Prior infection?** | No | No | Yes | Yes |
| **Number of adenovirus-vaccine doses:** | 1 | 2+ | 1 | 2+ |
| **Number of individuals:** | 4522 | 1489 | 1707 | 291 |
| **Median age (IQR):** | Median: 51<br>IQR: [36, 62] | Median: 64<br>IQR: [52, 69] | Median: 48<br>IQR: [36, 59] | Median: 61<br>IQR: [47, 69] |
| **Sex (counts):** | Female: 2356<br>Male: 2166<br>Unknown: 0 | Female: 719<br>Male: 770<br>Unknown: 0 | Female: 875<br>Male: 832<br>Unknown: 0 | Female: 129<br>Male: 162<br>Unknown: 0 |
| **Race (counts):** | White: 4227<br>Unknown: 13<br>Unable to Provide: <10<br>Black or African American: 118<br>Other: 70<br>Asian: 30<br>Chose Not to Disclose: 24<br>Native American and Pacific Islander: 25<br>Asian - Far East: 16<br>Asian - Indian Subcontinent: <10 | White: 1408<br>Unknown: <10<br>Unable to Provide: <10<br>Black or African American: 31<br>Other: 10<br>Asian: 10<br>Chose Not to Disclose: 11<br>Native American and Pacific Islander: <10<br>Asian - Far East: 10<br>Asian - Indian Subcontinent: <10 | White: 1593<br>Unknown: <10<br>Unable to Provide: <10<br>Black or African American: 38<br>Other: 36<br>Asian: 17<br>Chose Not to Disclose: <10<br>Native American and Pacific Islander: <10<br>Asian - Far East: <10<br>Asian - Indian Subcontinent: <10 | White: 277<br>Unknown: 0<br>Unable to Provide: 0<br>Black or African American: <10<br>Other: <10<br>Asian: <10<br>Chose Not to Disclose: 0<br>Native American and Pacific Islander: <10<br>Asian - Far East: <10<br>Asian - Indian Subcontinent: <10 |
| **Ethnicity (counts):** | Not Hispanic or Latino: 4139<br>Unknown: 27<br>Hispanic or Latino: 138<br>Chose Not to Disclose: 52 | Not Hispanic or Latino: 1440<br>Unknown: <10<br>Hispanic or Latino: 29<br>Chose Not to Disclose: 21 | Not Hispanic or Latino: 1611<br>Unknown: <10<br>Hispanic or Latino: 76<br>Chose Not to Disclose: 11 | Not Hispanic or Latino: 281<br>Unknown: 0<br>Hispanic or Latino: <10<br>Chose Not to Disclose: <10 |

### The rate of SARS-CoV-2 reinfections has increased during Omicron and increased duration since prior infection is associated with increased risk of reinfection

Next, we evaluated temporal trends in SARS-CoV-2 reinfections. Reinfection was defined by the record of a positive SARS-CoV-2 PCR test at least 60 days after a prior positive SARS-CoV-2 PCR test (with no positive tests during the intervening 60-day interval). We investigated patterns in the risk of reinfection with respect to the date of reinfection and the date of prior infection, over the entire study population regardless of vaccination status

To quantify the temporal patterns in incidence of reinfections, we calculated the relative risk of SARS-CoV-2 infection for previously infected individuals compared with a control cohort without previous infection, during each calendar month (**Figure 2a**). Reinfections were rare during the first year of the pandemic and remained rare during the Alpha and Delta variant surges in 2021 (**Figure 2a**, up to November 2021). However, the relative risk of infection in individuals with prior infections increased significantly during the Omicron surge (**Figure 2a**, December 2021 onward). For example, during January 2022, RR was 0.67 [0.63-0.71, 95% CI], while during October 2021 RR was 0.11 [0.10-0.14, 95% CI], indicating a ∼6-fold increase in the relative risk of reinfection.

**Figure 2:**
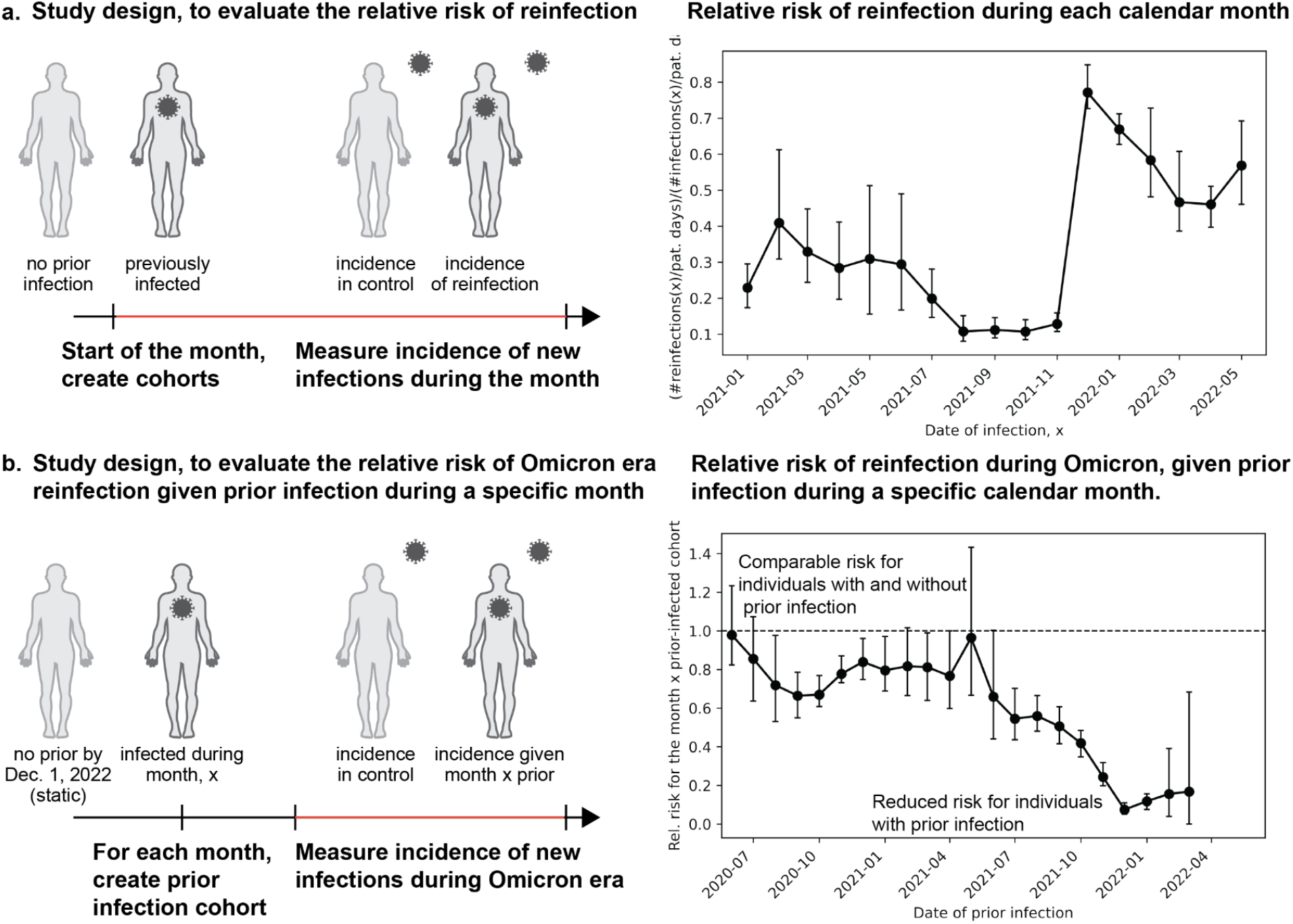
Risk of reinfection over time and relative to the date of prior infection. **(a)** Relative risk of reinfection during each calendar month, relative to the contemporary rate of first time infections. **(b)** Relative risk of reinfection during the Omicron era (December 1st, 2021 - May 29, 2022), for individuals with a prior infection during a specific calendar month (x-axis). The dashed horizontal guideline at y=1.0 indicates the expected result when the risk for individuals with a prior infection during the given calendar month (x-axis) is the same as for individuals without prior infection. Error bars indicate 95% confidence intervals.

To explore a possible relationship between the risk of reinfection during Omicron and the duration since prior SARS-CoV-2 infection, we evaluated the relative risk of reinfection during Omicron as a function of the month of prior infection (**Figure 2c**). Here, a relative risk of 1 indicates that an individual with a known prior infection is as likely to be infected during Omicron as an individual with no known prior infection. Individuals with prior infections between July, 2021, and November, 2021, had lower rates of reinfection during Omicron (e.g., RR = 0.24 [0.20-0.29, 95% CI] for prior infection during November, 2021). On the other hand, individuals with prior infections before June, 2021, had reinfections during Omicron at comparable rates with individuals that had no prior infection (RR = 0.80 [0.69-0.90] for prior infection during January, 2021).

### Rate of hospitalizations in patients with prior infection and vaccination is lower than in patients with vaccination only

Disease severity of Omicron infections was assessed by looking at the rates of hospitalization within 14 days of a positive PCR test, during the Omicron time-window (December 1, 2021 - May 29, 2022). Demographics in the resulting cohorts of SARS-CoV-2 infected individuals were similar between individuals with and without prior infection (**Table 3**). Overall, hospitalization rates for mRNA vaccinated individuals with a prior SARS-CoV-2 infection were lower than those for individuals with only mRNA vaccination (**Figure 3**). Specifically, the 14-day hospitalization rate in mRNA vaccinated patients with prior infection was lower than in those without prior infection (0.91% [0.34-1.6, 95% CI] versus 1.4% [1.1-1.7, 95% CI]). Consistently, we further observed that in patients with unknown vaccination status, those with prior infection had lower hospitalization rates than those without prior infection (1.7% [1.1-2.3, 95% CI] versus 3.3% [2.9-3.7], 95% CI) (**Figure S2**). We observed no statistically significant difference for the observed rate of hospitalization in mRNA-boosted patients with prior infection (1.9% [0.81-3.2, 95% CI]) compared to those without prior infection (1.8% [1.5-2.1, 95% CI]). We also observed increased rates of hospitalization in mRNA-boosted patients compared to mRNA-vaccinated patients, this result is expected given the prioritization of booster doses for individuals that are at risk of severe disease (**Table 3**).^13–15^

**Table 3:**
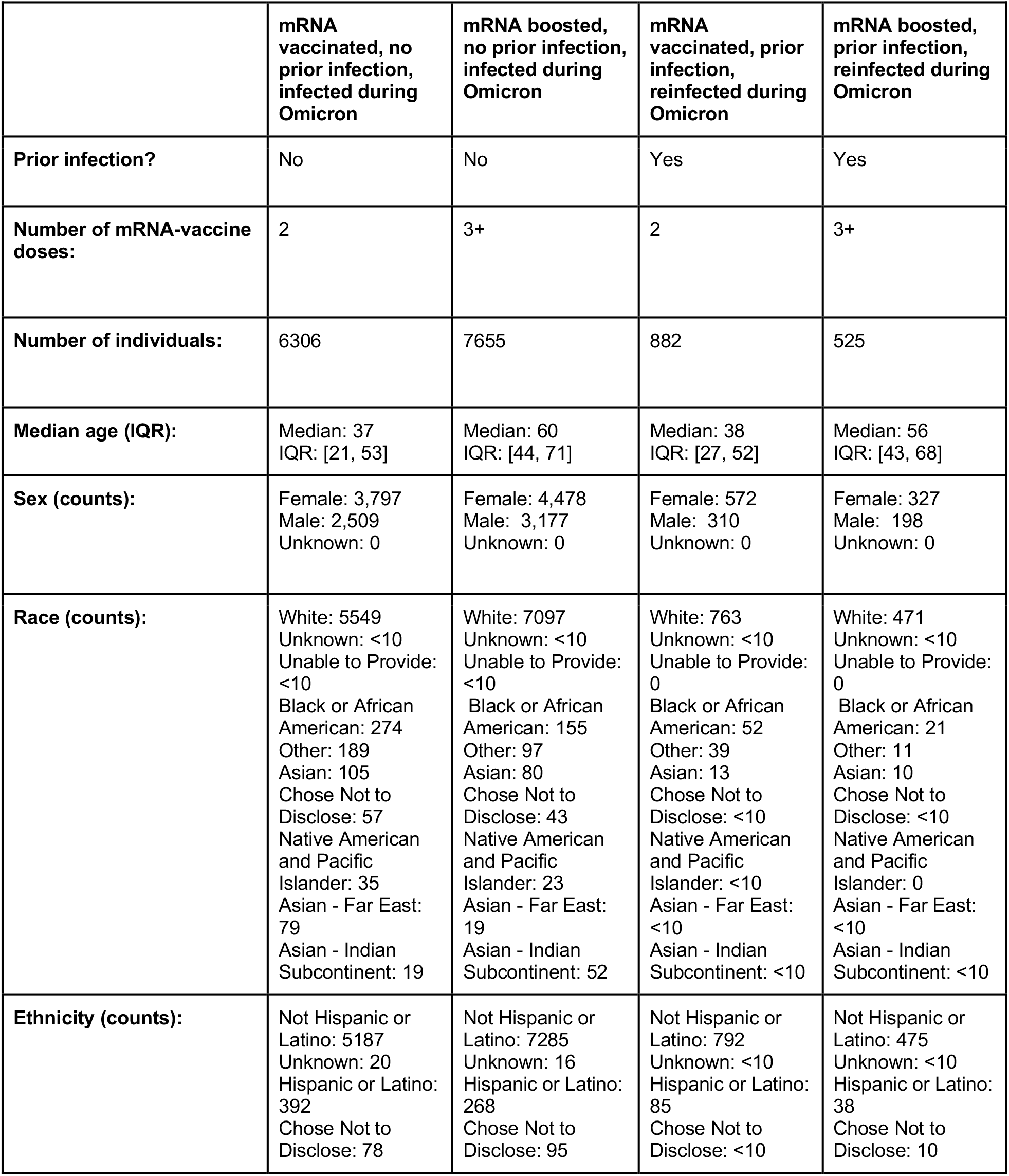
Cohort characteristics for mRNA-vaccinated, Omicron infected individuals.

**Figure 3:**
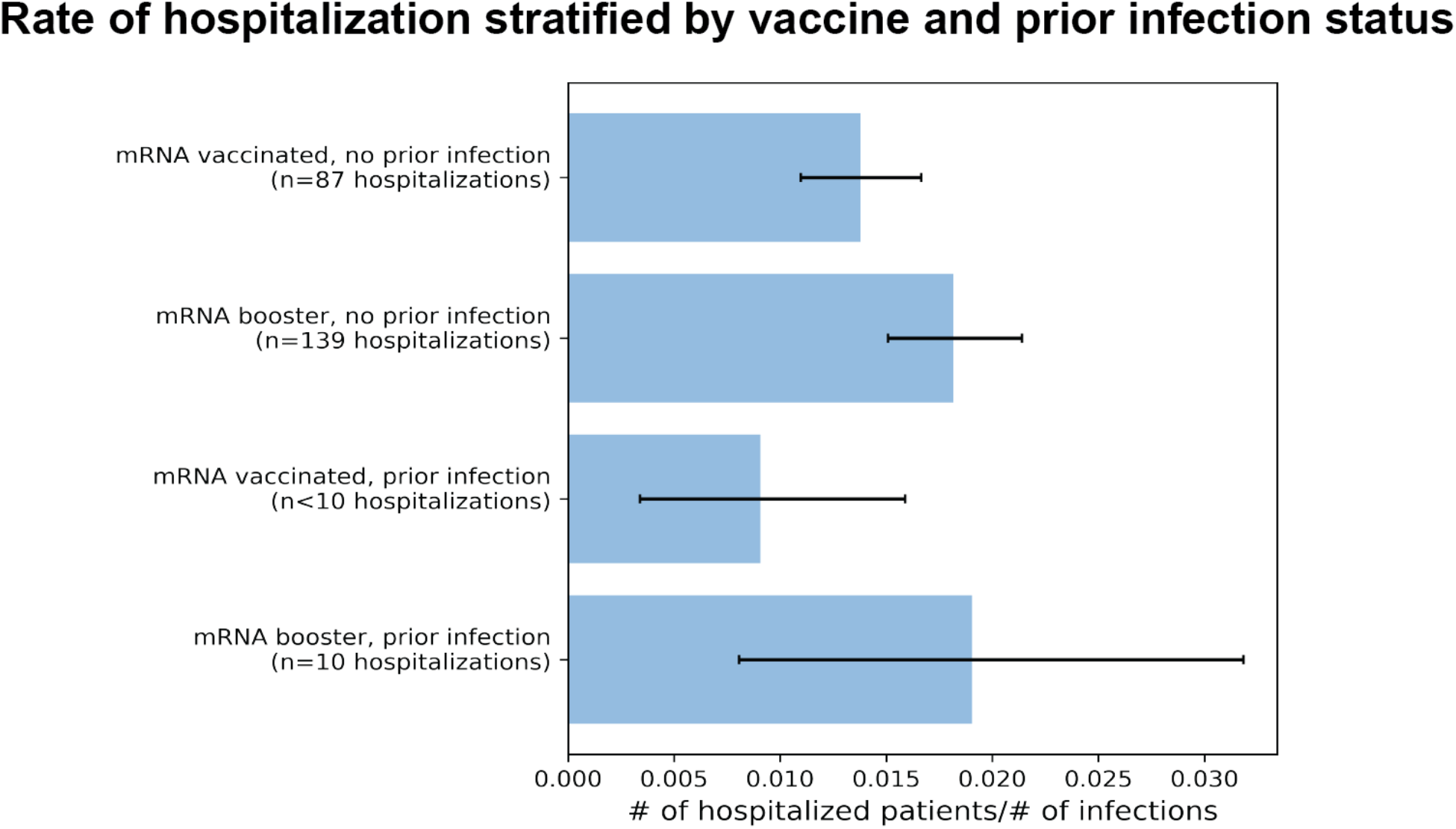
Hospitalization rate of COVID-19 patients during Omicron (December 2021 - June 2022), stratified by patient vaccination status and prior exposure to SARS-CoV-2. Shown is the fraction of infected individuals that were subsequently hospitalized, within 14 days of their positive PCR test. Error bars indicate 95% confidence intervals.

## Discussion

Even as individuals acquire immunity from prior infections, vaccinations and both, the risk of infection with and hospitalization from an evolving SARS-CoV-2 virus is still incompletely understood. Consistent with prior studies,^16,17^ our findings suggest that hybrid immunity or booster vaccination provides more protection against Omicron infections than full vaccination alone. Additionally, we show a drastic increase in SARS-CoV-2 reinfections during times of Omicron prevalence, consistent with prior studies in South Africa^5^ and the United States.^18^ Importantly, we find that the risk of reinfection is increased for patients with greater duration since their prior SARS-CoV-2 infection, providing evidence for a reduction of protection due to prior infection. In addition to reducing the risk of Omicron infection we also show that individuals with hybrid immunity are at lower risk of hospitalization due to an Omicron infection.

There are a few caveats to this study. First, only PCR tests and vaccinations that were recorded in the EHRs were considered when deriving the cohorts. We were unable to account for any externally administered vaccinations or tests (e.g. at-home rapid antigen tests) that were not also reported in the EHR. To reduce the impact of this limitation, we restricted our study population to those individuals with regular health encounters and a principal care provider at one of the study sites. Second, social behavioral aspects and health policy related factors such as social distancing and wearing of masks, which varied during the course of the pandemic, were not accounted for in this study. Third, there are individuals that have received more than one booster dose and understanding their level of protection is also warranted, given a recent study suggesting higher protection against infection among individuals that received four doses.^19^ In the present study, all individuals with booster doses are grouped together, regardless of the number of additional doses they received. Finally, during the process of EHR deidentification, there is a random time-shift of up to one month applied uniformly to all encounters for a given patient. As such, any results reported by date will effectively be a rolling average across data within a two-month timespan around that date.

Our findings regarding infection and hospitalization rates during the Omicron era further motivate complementary seroprevalence studies^10^ to gain a comprehensive understanding of the immunity conferred by combinations of vaccines, boosters, and prior infections. As new variant-targeted vaccine formulations are being developed such as Omicron-adapted boosters^20^ and the bivalent boosters^21^, accounting for protection conferred from prior infections, particularly prior Omicron infections, can better guide public health policy decisions around administering booster doses. Our study also motivates future work that specifically assesses the risk of Omicron reinfections from newly emerging subvariants like BA.5 and BA.2.12.1 in individuals previously infected with other Omicron sublineages like BA.1 or BA.2. This is especially relevant given early indications that the BA.5 and BA.2.12.1 Omicron subvariants can evade immunity acquired from prior infection with earlier Omicron variants.^20,22–25^

## Methods

### De-identified EHR data

For this analysis, we consider data from the nference Analytics Platform.^12^ This database includes de-identified HIPAA-compliant Electronic Health Records (EHRs) from over 6 million patients treated within US health systems.

The de-identification process, which went through the expert determination path allowed under the Health Insurance Portability and Accountability Act of 1996 (HIPAA) Privacy Rule de-identification standard, follows a “hiding in plain sight” approach to detect and obfuscate personal identifiers (e.g names, organizations, locations etc.) in free form text in the EHR with suitable surrogates.^12^ Identifiers in the structured database component of the EHR are either redacted (e.g names, addresses, etc.) or hashed using a one-way function with a secret salt (e.g patient IDs, to allow for collating different longitudinal information elements like diagnoses, medications, vaccinations, hospitalization, lab measurements etc. for a patient). Dates are time shifted randomly across patients, but uniformly across all events for a patient. Patient ages above 89 are grouped together. Only content previously analyzed for the presence of personal identifiers is allowed to pass through; so information elements not yet analyzed and approved, like newly introduced diagnosis codes, procedures or measurements may be filtered. Quasi-identifiers are generalized (e.g “Korean” ethnicity mapped to “Far East Asian”)

The nference Analytics Platform operates in a “data under glass” mode. Access to data is login protected and within the platform. Only aggregated insights are exported outside of the platform.

### Study population

The study population consists of all individuals that satisfy the following criteria: (1) have at least one SARS-CoV-2 PCR test before the study start date, December 1, 2021, (2) have at least one encounter per year on file, for the past two years, and (3) have a Principal Care Provider (PCP) within the study health systems on record. The last two criteria restrict our study population to those who are regularly receiving care within study health systems. These were put in place to ensure completeness of SARS-CoV-2 PCR testing data for individuals included in the analysis.

In order to assess Omicron-associate infection and hospitalization rates in groups with varying modalities of immunity, these patients were further divided into ten cohorts based on their vaccination and prior SARS-CoV-2 infection status, evaluated on December 1st, 2021:

Cohort 1 - No known vaccine, no prior: No COVID-19 vaccine on record, no prior positive SARS-CoV-2 test.

Cohort 2 - No known vaccine, prior: No COVID-19 vaccine on record, at least one prior positive SARS-CoV-2 test.

Cohort 3 - mRNA vaccinated, no prior: 2-dose mRNA vaccinated (BNT162b2 or mRNA-1273), no prior positive SARS-CoV-2 test.

Cohort 4 - mRNA booster, no prior: 3-dose+ mRNA vaccinated (BNT162b2 or mRNA-1273), no prior positive SARS-CoV-2 test.

Cohort 5 - mRNA vaccinated, prior: 2-dose mRNA vaccinated (BNT162b2 or mRNA-1273), at least one prior positive SARS-CoV-2 test.

Cohort 6 - mRNA booster, prior: 3-dose+ mRNA vaccinated (BNT162b2 or mRNA-1273), at least one prior positive SARS-CoV-2 test.

Cohort 7 - Adenovirus vaccinated, no prior: 1-dose adenovirus vaccinated (Ad26.CoV2.S), no prior positive SARS-CoV-2 test.

Cohort 8 - Adenovirus booster, no prior: 2-dose+ adenovirus vaccinated (Ad26.CoV2.S), no prior positive SARS-CoV-2 test.

Cohort 9 - Adenovirus vaccinated, prior: 1-dose adenovirus vaccinated (Ad26.CoV2.S), at least one prior positive SARS-CoV-2 test.

Cohort 10 - Adenovirus booster, prior: 2-dose+ adenovirus vaccinated (Ad26.CoV2.S), at least one prior positive SARS-CoV-2 test.

Here, a prior infection is defined as an infection confirmed by a positive SARS-CoV-2 PCR test before the study period start date, December 1, 2021. Cohort characteristics are provided in **Table 1** and **Table 2**.

### Calculation of eligible patient days for calculation of SARS-CoV-2 infection rates

Each individual included in the study contributes patient days between the study start date (December 1, 2021) and the study end date (May 29, 2022). The number of patient days contributed by an individual can be reduced based on the following censoring events:

#### Right censoring

Individuals were right censored upon (1) receiving any additional COVID-19 vaccine dose or (2) a positive SARS-CoV-2 PCR test, with no other positive test during the preceding 60 days.

#### Left censoring

Individuals were left censored until the first date where they had no positive SARS-CoV-2 PCR test on file during the preceding 60 days.

### Quantification of SARS-CoV-2 infections and reinfections

The following criteria are used to define incidence of SARS-CoV-2 infection for an individual, all criteria need to be satisfied: (1) a positive SARS-CoV-2 PCR test (2) no other positive test during the 60 days before the considered positive test. Here, the 60 day time period allows for the counting of SARS-CoV-2 reinfections while ensuring that consecutive positive tests are not due to a single persistent infection. A reinfection needs to satisfy the same criteria as those for an infection, with the additional criterion that the individual has a prior positive SARS-CoV-2 PCR test.

To quantify the risk of infections and reinfections, we report the number of eligible (re)infections during specified time-windows normalized by the number of eligible patient days contributed by the cohort.

We calculated 95% confidence intervals by performing bootstrapping with 1,000 samples. Specifically, sample cohorts of the same size as the original cohort were generated by sampling with replacement from the original cohort population, and 95% confidence intervals were determined using the percentile method.

### Quantification of the hospitalization rate following SARS-CoV-2 infection

A hospitalization associated with an Omicron infection is defined as one that occurs within 14 days of an individual’s first positive SARS-CoV-2 PCR test. The rate of hospitalization is calculated by dividing the number of hospitalized patients in a cohort by the total number of patients with Omicron infections in each cohort.

We calculated 95% confidence intervals by performing bootstrapping with 1,000 samples. Specifically, sample cohorts of the same size as the original cohort were generated by sampling with replacement from the original cohort population, and 95% confidence intervals were determined using the percentile method.

## Data Availability

This study involves analysis of de-identified Electronic Health Record (EHR) data via the nference Analytics Platform. Data shown and reported in this manuscript has been extracted from this environment using an established protocol for data extraction, aimed at preserving patient privacy. The data has been de-identified pursuant to an expert determination in accordance with the HIPAA Privacy Rule. Any data beyond what is reported in the manuscript, including but not limited to the raw EHR data, cannot be shared or released due the parameters of the expert determination to maintain the data de-identification. Contact corresponding authors for additional details regarding the nference nsights Analytics Platform.

## Data availability statement

This study involves analysis of de-identified Electronic Health Record (EHR) data via the nference Analytics Platform. Data shown and reported in this manuscript has been extracted from this environment using an established protocol for data extraction, aimed at preserving patient privacy. The data has been de-identified pursuant to an expert determination in accordance with the HIPAA Privacy Rule. Any data beyond what is reported in the manuscript, including but not limited to the raw EHR data, cannot be shared or released due the parameters of the expert determination to maintain the data de-identification. Contact corresponding authors for additional details regarding the nference *nsights* Analytics Platform.

## Supplementary Material

**Figure S1:**
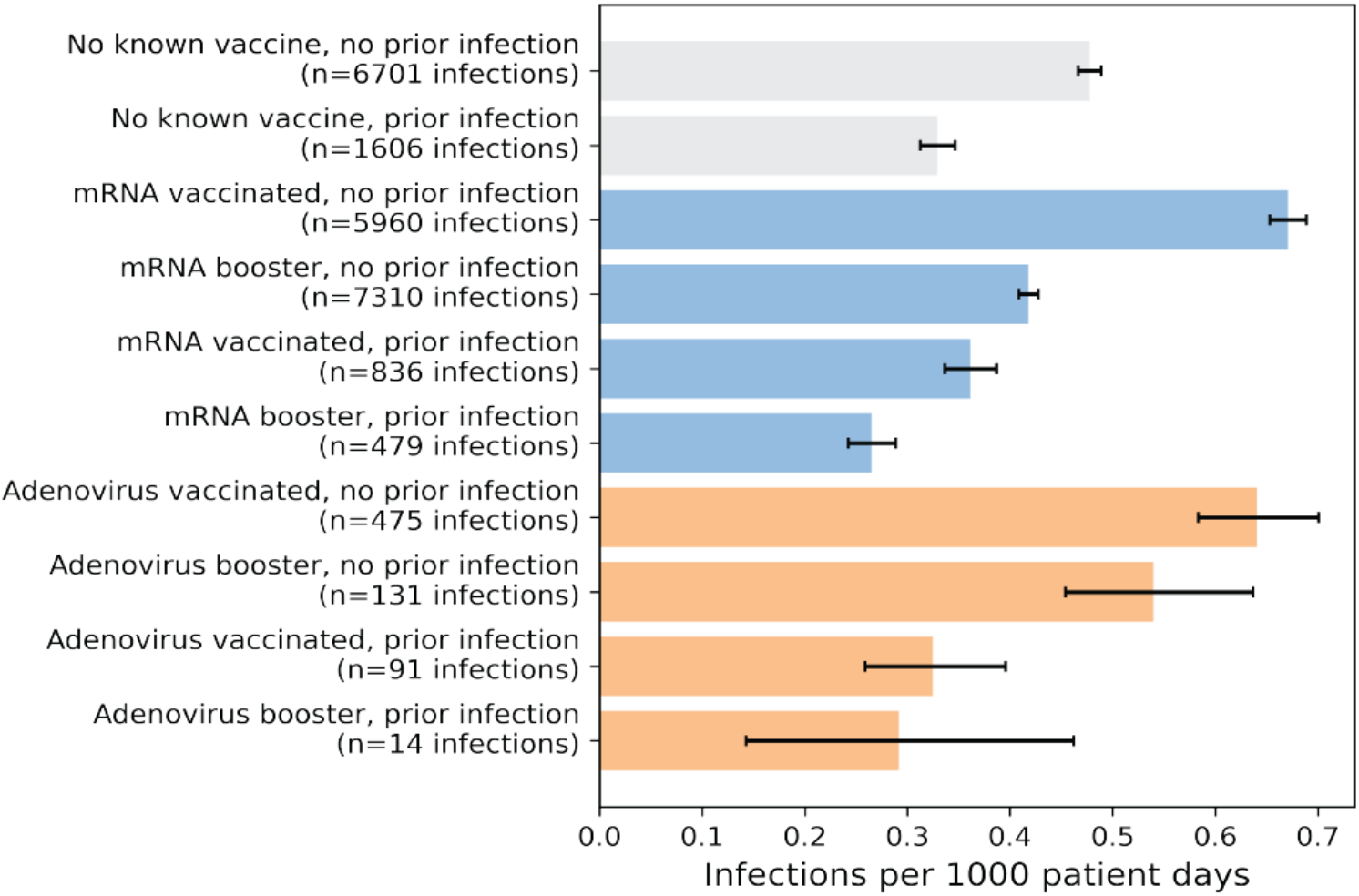
Rate of infections during Omicron (December 1, 2021 - June 2022), stratified by vaccination status and prior infection with SARS-CoV-2. The “no known vaccine” cohorts included here correspond to individuals with no vaccine information on file. These will be a mix of individuals with no vaccination or with varying degrees of undocumented vaccination. Error bars indicate 95% confidence intervals.

**Figure S2:**
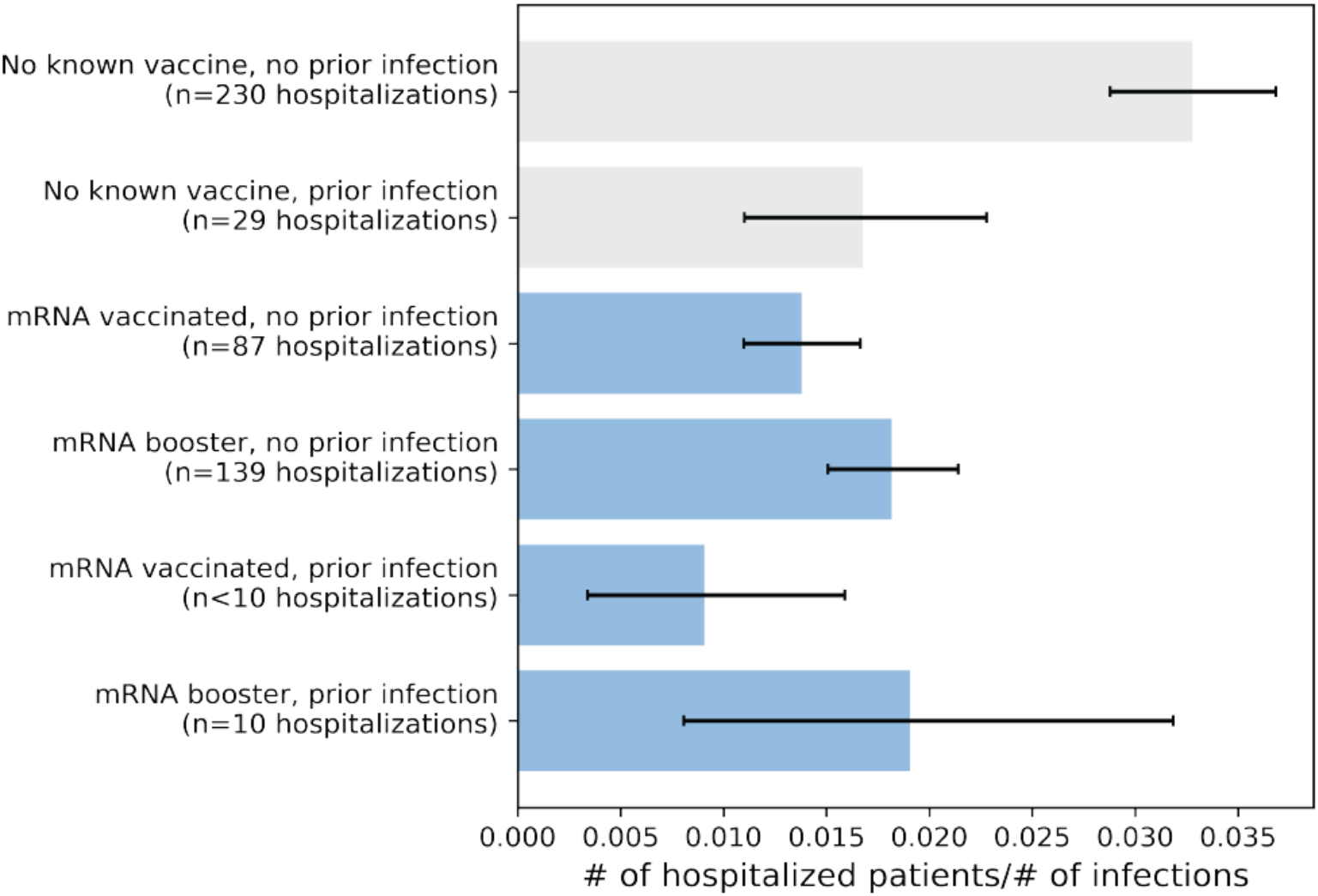
Hospitalization rate during Omicron (December 2021 - June 2022), stratified by patient vaccination status and prior exposure to SARS-CoV-2. The “no known vaccine” cohorts included here correspond to individuals with no vaccine information on file. Error bars indicate 95% confidence intervals.

## Notes

### Competing Interest Statement

MN, MJMN, PL, VA, JR, SA, AV, and VS are employees of nference and have financial interests in the company. nference collaborates with health systems, bio-pharmaceutical companies, and academic medical centers on data science initiatives unrelated to this study. These collaborations had no role in the study design, data collection, and analysis, decision to publish, or preparation of the manuscript.

### Funding Statement

This study did not receive any external funding

### Summary of Updates

Minor updates to the manuscript text.

## References

1. COVID-19 Map. Johns Hopkins Coronavirus Resource Centerhttps://coronavirus.jhu.edu/map.html.

2. Murugadoss, K. et al. Continuous genomic diversification of long polynucleotide fragments drives the emergence of new SARS-CoV-2 variants of concern. PNAS Nexus 1, (2022).

3. McCarthy, K. R. et al. Recurrent deletions in the SARS-CoV-2 spike glycoprotein drive antibody escape. Science 371, 1139–1142 (2021).

4. Kemp, S. A. et al. SARS-CoV-2 evolution during treatment of chronic infection. Nature 592, 277–282 (2021).

5. Pulliam, J. R. C. et al. Increased risk of SARS-CoV-2 reinfection associated with emergence of Omicron in South Africa. Science 376, eabn4947 (2022).

6. Classification of Omicron (B.1.1.529): SARS-CoV-2 Variant of Concern. https://www.who.int/news/item/26-11-2021-classification-of-omicron-(b.1.1.529)-sars-cov-2-variant-of-concern.

7. COVID-19 Map. Johns Hopkins Coronavirus Resource Center https://coronavirus.jhu.edu/map.html.

8. Puranik, A. et al. Durability analysis of the highly effective BNT162b2 vaccine against COVID-19. bioRxiv (2021) doi:10.1101/2021.09.04.21263115.

9. Puranik, A. et al. Durability analysis of the highly effective mRNA-1273 vaccine against COVID-19. PNAS Nexus 1, (2022).

10. Interim statement on hybrid immunity and increasing population seroprevalence rates. https://www.who.int/news/item/01-06-2022-interim-statement-on-hybrid-immunity-and-increasing-population-seroprevalence-rates.

11. Israel, A. et al. Elapsed time since BNT162b2 vaccine and risk of SARS-CoV-2 infection: test negative design study. BMJ 375, e067873 (2021).

12. Murugadoss, K. et al. Building a best-in-class automated de-identification tool for electronic health records through ensemble learning. Patterns (N Y) 2, 100255 (2021).

13. Arbel, R. et al. BNT162b2 Vaccine Booster and Mortality Due to Covid-19. N. Engl. J. Med. 385, 2413–2420 (2021).

14. Bar-On, Y. M. et al. Protection of BNT162b2 Vaccine Booster against Covid-19 in Israel. N. Engl. J. Med. 385, 1393–1400 (2021).

15. CDC. COVID-19 Vaccine Booster Shots. https://www.cdc.gov/coronavirus/2019-ncov/vaccines/booster-shot.html?s_cid=11706:cdc%20covid%20booster:sem.ga:p:RG:GM:gen:PTN:FY22 (2021).

16. Altarawneh, H. N. et al. Effects of Previous Infection and Vaccination on Symptomatic Omicron Infections. N. Engl. J. Med. (2022) doi:10.1056/NEJMoa2203965.

17. Bates, T. A. et al. Vaccination before or after SARS-CoV-2 infection leads to robust humoral response and antibodies that effectively neutralize variants. Sci Immunol 7, eabn8014 (2022).

18. COVID-19 Reinfection Data. Department of Health https://coronavirus.health.ny.gov/covid-19-reinfection-data.

19. Bar-On, Y. M. et al. Protection by a Fourth Dose of BNT162b2 against Omicron in Israel. N. Engl. J. Med. (2022) doi:10.1056/NEJMoa2201570.

20. Board Members. Pfizer and BioNTech Announce Omicron-Adapted COVID-19 Vaccine Candidates Demonstrate High Immune Response Against Omicron. https://www.pfizer.com/news/press-release/press-release-detail/pfizer-and-biontech-announce-omicron-adapted-covid-19.

21. Moderna Announces Omicron-Containing Bivalent Booster Candidate mRNA-1273.214 Demonstrates Superior Antibody Response Against Omicron. https://investors.modernatx.com/news/news-details/2022/Moderna-Announces-Omicron-Containing-Bivalent-Booster-Candidate-mRNA-1273.214-Demonstrates-Superior-Antibody-Response-Against-Omicron/default.aspx.

22. Cao, Y. et al. BA.2.12.1, BA.4 and BA.5 escape antibodies elicited by Omicron infection. Nature 1–3 (2022).

23. Tuekprakhon, A. et al. Antibody escape of SARS-CoV-2 Omicron BA.4 and BA.5 from vaccine and BA.1 serum. Cell 185, 2422–2433.e13 (2022).

24. Hachmann, N. P. et al. Neutralization Escape by SARS-CoV-2 Omicron Subvariants BA.2.12.1, BA.4, and BA.5. N. Engl. J. Med. (2022) doi:10.1056/NEJMc2206576.

25. CDC. COVID Data Tracker. https://covid.cdc.gov/covid-data-tracker/#datatracker-home.

